# Hypergraph-Based Doubly Robust Estimation for Causal Inference of Drug Combination Effects in Heart Failure Treatment

**DOI:** 10.64898/2025.12.11.25342026

**Authors:** Tingsong Xiao, Ya-Yun Yeh, Yao An Lee, Yi Guo, Jingchuan Guo, Zhe Jiang

**Affiliations:** Department of Department of Computer and Information Science and Engineering, University of Florida, Gainesville, FL, USA; Department of Pharmacy Practice, Purdue University, Indianapolis, IN, USA; Department of Health Outcomes and Biomedical Informatics, University of Florida, Gainesville, FL, USA

## Abstract

Disease management for heart failure with preserved ejection fraction (HFpEF) requires understanding the comparative effectiveness of real-world drug combinations rather than single agents. Standard randomized controlled trials (RCTs) for multi-drug regimens are prohibitively expensive, slow, and often infeasible at scale, motivating the use of causal machine learning methods on large-scale electronic health records (EHRs). However, reliable estimation of treatment effects is challenging due to the high-order drug–drug interactions, strong confounding factors, and patient heterogeneity across sex, age, and other statuses. Existing causal machine learning methods mostly focus on comparing treatments with pairwise drug combinations. Techniques for multi-drug regimens are less studied. To fill this gap, we propose a **H**ypergraph–based **D**oubly **R**obust framework for HFpEF (**HyperDR**), which represents six HF drug classes as nodes and observed multi-drug regimens as hyperedges, and uses a hypergraph neural network to learn shared representations for both drugs and combinations from cross-sectional EHR data. On top of these representations, we jointly train a propensity-score model and an outcome model with a doubly robust objective that combines cross-entropy losses with an augmented inverse-probability–weighted regularizer. This enables consistent treatment-effect estimation when either component is correctly specified while stabilizing learning under rare regimens. Experiments on a real-world HFpEF cohort show that HyperDR improves outcome prediction (hospitalization risk) compared with baseline methods. We also did case studies to interpret model results in treatment rankings across different patient subgroups.

## 1 Introduction

Heart failure with preserved ejection fraction (HFpEF) accounts for approximately 50% of all heart failure cases, and its prevalence is increasing due to aging populations and comorbidities (Tsao et al., 2023; Redfield, 2016). Unlike heart failure with reduced ejection fraction (HFrEF), for which large-scale randomized controlled trials (RCTs) have already established clear multi-drug guideline-directed medical therapies (GDMT) (Heidenreich et al., 2022; McMurray and Packer, 2021), evidence for optimal combination strategies in HFpEF is less established. While sodium-glucose cotransporter-2 (SGLT2) inhibitors are now a good recommendation (Solomon et al., 2022), HFpEF management in real-world practice involves complex polypharmacy. Because HFpEF patients often have multiple comorbidities, such as hypertension, obesity, and diabetes, clinicians routinely prescribe combinations of SGLT2 inhibitors, renin-angiotensin-aldosterone system (RAAS) inhibitors, beta-blockers, and diuretics.

Identifying the most effective combination for an individual patient is difficult. RCTs cannot feasibly test every possible combination of three or four drug classes (Lienhardt et al., 2011). Additionally, RCTs often exclude patients with severe multimorbidity, limiting their applicability to the broader patient population. Observational real-world data (RWD) provide a large sample size for analysis but presents two main challenges: confounding by indication (where treatment assignment depends on patient severity) and high dimensionality (modeling many possible drug combinations) (Hernán and Robins, 2016).

Causal deep learning (CDL) has recently emerged as a method to estimate treatment effects from observational data. Techniques like representation learning can balance covariates between treatment groups to reduce confounding (Johansson et al., 2016; Shi et al., 2019). Recent studies have applied these methods to rank treatments for diabetes (Belthangady et al., 2022) and hypertension (Liu et al., 2025). However, most existing models represent drug combinations as simple lists or sequences. This approach fails to capture the high-order interactions within a drug regimen, where the combined effect of multiple drugs may differ from the sum of their individual effects.

In this study, we fill this gap by developing a hypergraph-based, doubly robust causal deep learning framework for estimating the effects of HFpEF drug combinations on subsequent HFpEF hospitalization. We represent six key drug categories, including sacubitril/valsartan, SGLT2 inhibitors, loop diuretics, Mineralocorticoid Receptor Antagonists (MRAs), RAAS inhibition, and *β*-blockers, as nodes in a drug hypergraph, and represent each observed multi-drug regimen as a hyperedge. A hypergraph neural network learns shared embeddings for drug nodes and induced drug combination (hyperedge) representations. These embeddings are then used jointly with patient covariates to parameterize two coupled modules: (i) a propensity-score predictor that models the assignment probability of each drug combination treatment, and (ii) an outcome predictor that estimates the risk of HFpEF rehospitalization under each potential combination. To integrate these components, we design a doubly robust training objective that links the two modules, providing treatment-effect estimates that remain consistent when either model is correctly specified, while simultaneously promoting covariate balance and stabilizing estimation under infrequent treatment combinations. Unlike prior causal deep learning models that treat each treatment regimen as an unstructured label, our hypergraph formulation shares information across related combinations and encodes higher-order drug–drug interactions. We then apply this framework to a large HFpEF EHR cohort to (1) rank drug combinations by estimated causal effect on rehospitalization, (2) characterize patient heterogeneity in optimal combinations across different clinically relevant subgroups, and (3) compare our approach with both deep causal and non-deep baselines. Our results provide both methodological advances, i.e., a hypergraph-based doubly robust estimator tailored to multi-drug therapy, and clinically interpretable insights into HFpEF pharmacotherapy that potentially inform future guidelines, trial design, and decision support systems.

## 2 Related Work

### 2.1 Causal Inference in Observational Studies

The fundamental challenge in observational studies lies in estimating counterfactual outcomes while rigorously adjusting for confounding. Standard statistical methods, such as Inverse Probability of Treatment Weighting (IPTW), often suffer from instability in high-dimensional settings due to insufficient overlap in propensity scores among treatment groups (Imbens and Rubin, 2015). Deep representation learning offers a robust alternative by mapping patient covariates into a latent space where the distributions of treated and control groups are balanced. Seminal architectures like TAR-Net (Shalit et al., 2017) and DragonNet (Shi et al., 2019) utilize neural networks to learn these balanced representations; DragonNet, in particular, employs a regularization term to ensure learned features are predictive of both treatment assignment and outcomes, a principle we adopt in our framework.

Extending these methodologies from binary interventions to multiple treatment combinations presents additional complexity. Recent approaches have attempted to adapt deep learning for this purpose: TransTEE (Zhang et al., 2024) employs transformer models to handle continuous or structured treatments, while NCORE (Parbhoo et al., 2021) explicitly models interactions between different treatment arms. Most relevantly, the METO framework introduced a multi-treatment encoding mechanism to handle drug sequences for hypertension (Liu et al., 2025). However, these methods largely treat combinations as discrete categories or sequential lists. They generally fail to explicitly model the set-based structure inherent in polypharmacy, limiting their ability to fully capture the complex, high-order interactions within multi-drug regimens.

## 3 Method

### 3.1 Input Representation

Our basic unit of analysis is a *snapshot* of a patient’s health state and treatment. Each patient’s longitudinal EHR history is first split into a series of temporal snapshots (e.g., merged 100-day windows). We then pool all snapshots from all patients together and treat them as observational data for model training and effect estimation.

#### Snapshot-level observational dataset

We denote by *N* the total number of snapshots (across all patients), and by

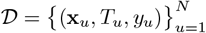

the resulting observational dataset, where each index *u* refers to the *u*-th snapshot. For each snapshot *u*:

- **x**_*u*_ ∈ ℝ ^*d*^ is a covariate vector summarizing the patient’s state at the snapshot (demographics, comorbidities, labs, vitals, etc.).
- *T*_*u*_ ⊆ {1, … , *K*_drug_} is the treatment combination prescribed during the snapshot, encoded as a subset of HF-relevant drug classes, where *K*_drug_ = 6 corresponds to sacubitril/valsartan, SGLT2 inhibitors, loop diuretics, MRAs, RAAS inhibitors, and *β*-blockers.
- *y*_*u*_ ∈ { 0, 1} is a binary outcome indicating whether the patient experiences HFpEF hospitalization in the follow-up period associated with this snapshot (e.g., by the next encounter).

For notational simplicity, in the following we omit the snapshot index *u* whenever there is no ambiguity and write (**x**, *T, y*).

#### Predefined drug hypergraph

To capture higher-order pharmacologic relationships among drug classes and treatment combinations, we construct a predefined drug hypergraph

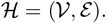

The node set

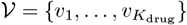

contains one node for each drug class. We scan all snapshots in 𝒟 and collect all unique combination patterns *T* ⊆ {1, … , *K*_drug_}. Each distinct pattern (e.g., {SGLT2 inhibitor + loop diuretic + *β*-blocker}) is mapped to a hyperedge *e* ∈ ℰ that connects exactly the corresponding drug-class nodes:

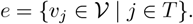

Thus, every snapshot-level treatment *T* is associated with a hyperedge *e* ∈ ℰ, and the collection of all such hyperedges defines the drug hypergraph ℋ, which we use to learn shared representations for drug classes and combination-level regimens.

### 3.2 Hypergraph-Based Drug and Treatment Representation

Given ℋ = (𝒱, ℰ), we construct low-dimensional embeddings for both drug classes and treatment combinations.

#### Incidence structure

Let *K*_drug_ = |𝒱| and *K*_comb_ = |ℰ| denote the numbers of drug classes and distinct combinations, respectively. We define the incidence matrix

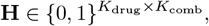

where **H**_*j,e*_ = 1 if drug-class node *v*_*j*_ participates in hyperedge *e*, and **H**_*j,e*_ = 0 otherwise. We also define diagonal degree matrices

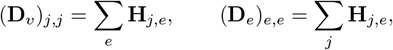

and a diagonal matrix of non-negative hyperedge weights

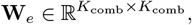

which is learned.

#### Drug-class embeddings

Let 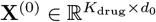 be the initial drug-node feature matrix. A hyper-graph neural network (HGNN) layer updates node embeddings via

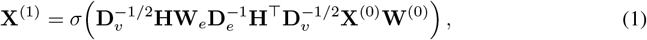

where 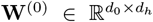 is a trainable weight matrix, *σ*(·) is a nonlinearity (e.g., ReLU), and 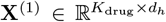 are updated node embeddings. Stacking *L* such layers yields the final drug-class embedding matrix

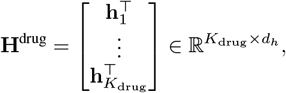

where 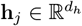 is the embedding of drug-class node *v*_*j*_.

#### Combination (hyperedge) embeddings

For each hyperedge *e* ∈ ℰ (i.e., each distinct treatment combination), we aggregate the embeddings of its constituent drug classes using a permutation-invariant function:

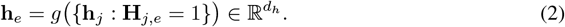

We use an attention-weighted average:

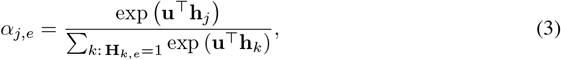

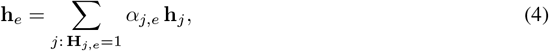

where 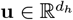 is a learnable query vector. Collecting all combination embeddings yields

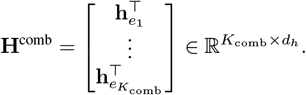

For a snapshot (**x**, *T, y*) ∈ 𝒟, the treatment set *T* corresponds to a hyperedge *e* ∈ ℰ, and we denote its embedding by

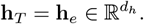

These combination-level embeddings are shared across all snapshots that use the same regimen, enabling information sharing across related treatment patterns.

### 3.3 Shared Representation and Doubly Robust Prediction Modules

Given snapshot-level covariates **x** and the corresponding treatment embedding **h**_*T*_ , our model learns a shared representation that feeds into two coupled prediction modules: a propensity-score model and an outcome model.

#### Shared snapshot–treatment representation

We first map covariates into a latent space via an MLP:

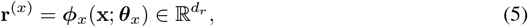

and then combine patient and treatment information via concatenation and a second encoder:

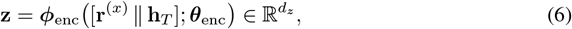

where **z** is a shared representation for the snapshot, and ***θ***_*x*_, ***θ***_enc_ are trainable parameters.

#### Propensity-score module

Let *K*_comb_ denote the number of distinct treatment combinations. We model the assignment probability of each combination *e*_*a*_ ∈ ℰ at a snapshot as

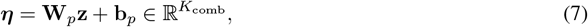

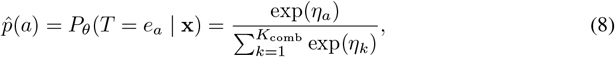

where 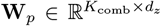 and 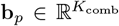 are trainable parameters, and *η*_*a*_ is the *a*-th entry of ***η***. For the observed treatment *T* (with index *a*^⋆^), the propensity loss is the standard multi-class cross-entropy

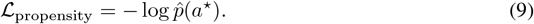

#### Outcome module

We model the conditional mean of the binary outcome under the observed treatment as

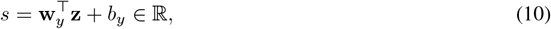

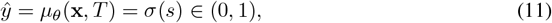

where 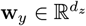 and *b*_*y*_ ∈ ℝ are trainable parameters, and *σ*(·) is the logistic function. The outcome loss is the standard binary cross-entropy

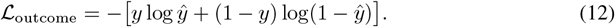

#### Doubly robust coupling via pseudo-outcomes

To couple the two modules and obtain doubly robust treatment-effect estimates, we construct an augmented inverse-probability–weighted (AIPW) pseudo-outcome for each combination *e*_*a*_:

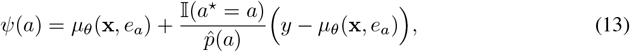

where 𝕀(·) is the indicator function and *µ*_*θ*_(**x**, *e*_*a*_) is obtained by replacing **h**_*T*_ with 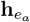 in the computation of **z**. Under standard causal assumptions, the empirical average of *ψ*(*a*) over snapshots is a doubly robust estimator of the marginal risk under combination *a*; it is consistent if either the propensity model 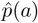 or the outcome model *µ*_*θ*_(**x**, *e*_*a*_) is correctly specified.

To stabilize learning and explicitly align the two heads, we introduce a residual-based regularizer:

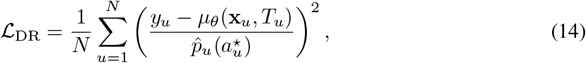

where we temporarily reintroduce the snapshot index *u* for clarity, and 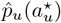 is the estimated propensity for the observed combination in snapshot *u*. This term penalizes large inverse-probability–weighted residuals, encouraging agreement between the propensity and outcome modules and improving the finite-sample stability of the AIPW estimator.

#### Total training objective

The overall loss combines factual prediction accuracy with the doubly robust regularization:

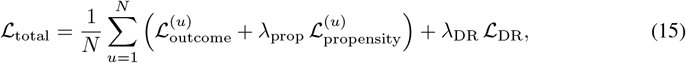

where 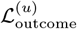 and 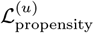 are the losses in Eq. 12–Eq. 9 evaluated at snapshot *u*, and *λ*_prop_, *λ*_DR_ ≥ 0 control the strength of the auxiliary terms. We optimize ℒ_total_ using stochastic gradient-based methods over mini-batches of snapshot-level samples.

### 3.4 Causal Effect Estimation and Treatment Ranking

After training, we use the learned model to estimate causal effects of treatment combinations and to construct treatment rankings.

#### Marginal risks and average treatment effects

For each combination *e*_*a*_ ∈ ℰ , we estimate the marginal risk of HFpEF hospitalization using the AIPW pseudo-outcome:

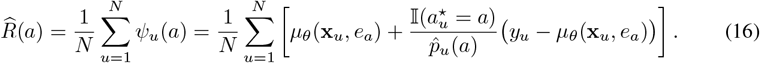

For any pair of combinations (*a, b*), the average treatment effect (ATE) is estimated by

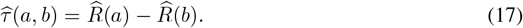

Subgroup-specific risks and ATEs (e.g., by sex, race, or obesity) are obtained by restricting the averages in Eq. 16 to snapshots belonging to the subgroup of interest.

#### Population-level and individualized rankings

At the population level, we rank treatment combinations by their estimated marginal risk 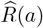, with lower values indicating more favorable regimens. For an individual patient profile 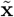 at decision time, we can form an individualized ranking by sorting combinations {*e*_*a*_} according to the predicted conditional risk 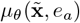, optionally combining these predictions with subgroup-level 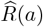 as a prior. Because treatment combinations are represented as hyperedges in a shared drug hypergraph and encoded by a common HGNN, these rankings exploit pharmacologic relationships and higher-order drug–drug interactions, while the doubly robust objective helps ensure that the implied treatment-effect estimates are robust to misspecification in either the propensity or outcome component.

## 4 Experiments

### 4.1 Prediction Performance of Propensity and Outcome Models

We first evaluate how well the proposed hypergraph-based framework predicts (1) treatment assignment (propensity scores) and (2) HFpEF hospitalization outcomes at the snapshot level.

Table 1 compares our HyperDR model with a multilayer perceptron (MLP) baseline that uses the same input covariates but treats each treatment combination as an unstructured label.

**Table 1:** Performance of propensity and outcome models on the HFpEF snapshot dataset. “Propen-sity” evaluates multi-class treatment assignment prediction; “Outcome” evaluates binary prediction of HFpEF hospitalization.

| Metric | MLP |  | HyperDR |  |
| --- | --- | --- | --- | --- |
|  | Propensity | Outcome | Propensity | Outcome |
| Accuracy | 0.22 | 0.89 | 0.23 | **0.90** |
| Precision | 0.23 | 0.67 | 0.23 | **0.76** |
| Recall | 0.23 | 0.68 | 0.23 | **0.72** |
| F1-score | 0.23 | 0.67 | 0.23 | **0.74** |
| AUC | – | 0.92 | – | **0.93** |

For the propensity task, both models achieve low but comparable performance (accuracy and F1-score around 0.22–0.23). This is expected given the large number of possible treatment combinations and strong overlap between regimens, and is consistent with prior work showing that prediction of multi-valued treatment assignment is difficult in real-world EHR data.

For the outcome task, HyperDR consistently outperforms the MLP across all metrics. Compared with MLP, the HyperDR model improves precision from 0.67 to 0.76, recall from 0.68 to 0.72, F1-score from 0.67 to 0.74, and AUC from 0.92 to 0.93. These gains indicate that explicitly modeling drug relations and higher-order combinations on the hypergraph leads to more informative treatment representations and better discrimination of HFpEF hospitalization risk, which is crucial for downstream treatment ranking and subgroup analyses.

### 4.2 Subgroup Analysis

In clinical practice, the effectiveness and safety of HFpEF treatments can differ across patient sub-groups defined by sex and age. To explore such heterogeneity, we stratify the snapshot-level cohort into clinically relevant subgroups and, within each stratum, estimate the marginal risk of HFpEF hospitalization for every observed treatment combination using the doubly robust procedure described in Section 3. For each subgroup, treatment combinations (hyperedges) are then ranked by their estimated average outcome risk (lower is better).

#### 4.2.1 Sex-stratified treatment ranking

We first consider sex as the stratification variable and divide the HFpEF snapshot cohort into male and female patients. Within each sex, we retain only treatment combinations that appear at least a minimum number of times in the training data to avoid unstable estimates for extremely rare regimens. For every remaining treatment hyperedge, we estimate its average probability of HFpEF hospitalization among snapshots belonging to that sex using the doubly robust procedure described in Section 3, and then rank all regimens in ascending order of estimated risk (lower is better).

For interpretability, Table 2 lists the mapping from numeric treatment IDs to specific combinations of the six HF drug classes (RAAS inhibitors, *β*-blockers, loop diuretics, MRAs, SGLT2 inhibitors, and sacubitril/valsartan). Figure 2 visualizes the resulting sex-specific rankings: the left panel shows the top-ranked regimens for male patients, and the right panel shows the corresponding ranking for female patients, with bar lengths indicating the estimated average hospitalization risk under each regimen.

**Table 2:** Mapping from treatment ID to drug-class combinations.

| Treatment ID | Treatment combination |
| --- | --- |
| 0 | RAAS |
| 1 | Beta_blockers |
| 2 | Beta_blockers, RAAS |
| 3 | Beta_blockers, Loop_diuretics, RAAS |
| 4 | Loop_diuretics |
| 5 | Beta_blockers, Loop_diuretics |
| 6 | Loop_diuretics, RAAS |
| 7 | MRAs, RAAS |
| 8 | MRAs |
| 9 | Beta_blockers, Loop_diuretics, MRAs, RAAS |
| 10 | Beta_blockers, MRAs, RAAS |
| 11 | Beta_blockers, MRAs |
| 12 | Beta_blockers, Loop_diuretics, MRAs |
| 13 | Loop_diuretics, MRAs |
| 14 | Loop_diuretics, MRAs, RAAS |
| 15 | SGLT2 |
| 16 | RAAS, SGLT2 |
| 17 | Beta_blockers, Loop_diuretics, RAAS, Sacubitril_Valsartan |
| 18 | Beta_blockers, Loop_diuretics, MRAs, RAAS, Sacubitril_Valsartan |
| 19 | Beta_blockers, RAAS, Sacubitril_Valsartan |
| 20 | RAAS, Sacubitril_Valsartan |
| 21 | Beta_blockers, Loop_diuretics, RAAS, SGLT2 |
| 22 | Beta_blockers, SGLT2 |
| 23 | MRAs, SGLT2 |
| 24 | Beta_blockers, MRAs, RAAS, SGLT2 |
| 25 | Loop_diuretics, SGLT2 |
| 26 | Beta_blockers, RAAS, SGLT2 |
| 27 | Loop_diuretics, RAAS, Sacubitril_Valsartan |

**Figure 1:**
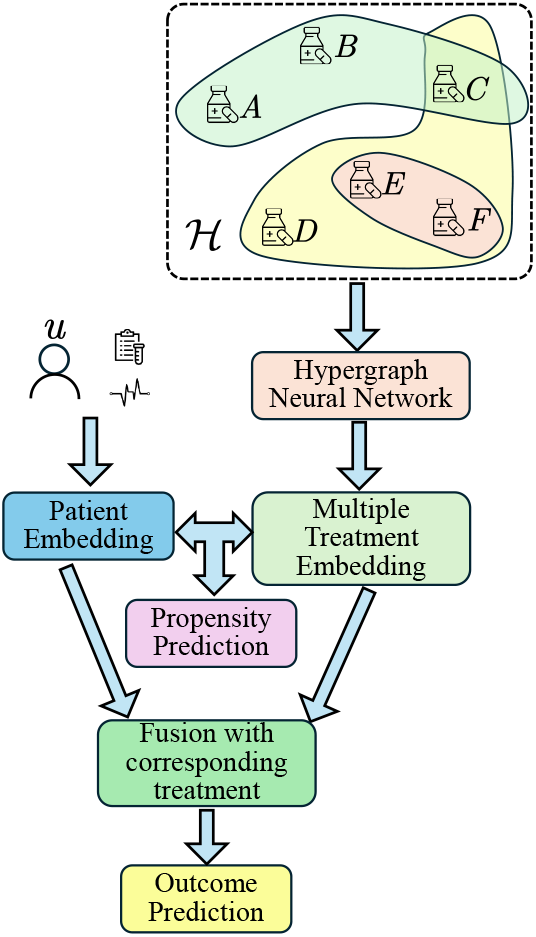
Overview of the HyperDR framework. The architecture integrates a hypergraph neural network for drug combinations with a patient encoder, jointly optimizing propensity and outcome prediction for doubly robust causal inference.

**Figure 2:**
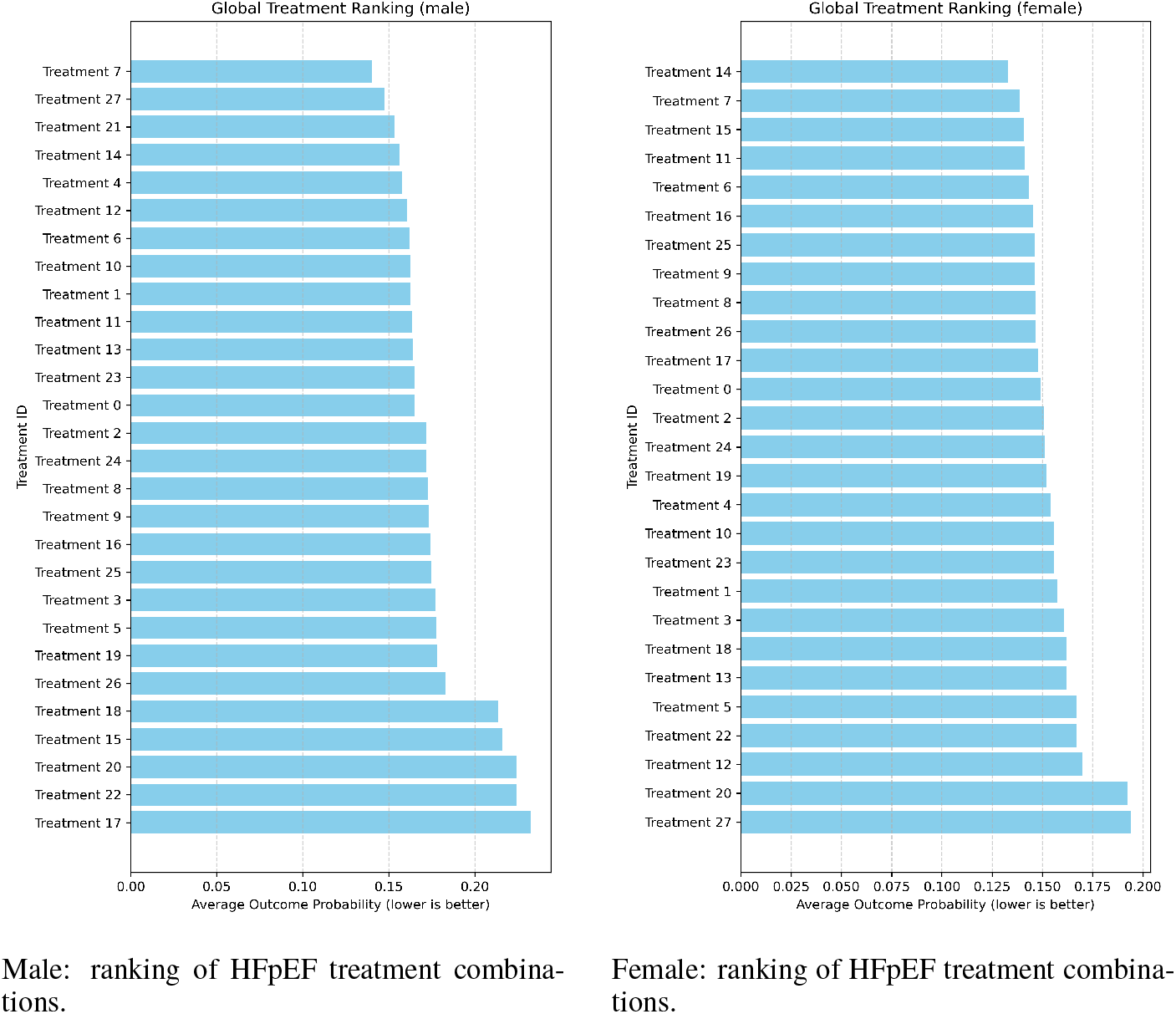
Sex-stratified ranking of HFpEF treatment combinations for male (left) and female (right) patients. Lower bars correspond to lower estimated HFpEF hospitalization risk.

Across both sexes, combination therapies that include RAAS inhibition together with one or more diuretics tend to occupy the top of the ranking, suggesting consistently lower HFpEF hospitalization risk compared with simpler regimens such as single-agent RAAS therapy. Several multi-drug combinations that incorporate newer agents (e.g., SGLT2 inhibitors or sacubitril/valsartan in addition to RAAS and diuretics) shift position between the male and female rankings, indicating potential sex-specific differences in comparative effectiveness.

#### 4.2.2 Age-stratified treatment ranking

We next stratify the cohort by age to examine whether the comparative effectiveness of treatment combinations differs between older and younger patients. Snapshots are divided into two groups based on age at the snapshot (*>* 65 years vs. ≤ 65 years). Within each age stratum, we keep only treatment combinations that occur at least a minimum number of times in the training data to reduce instability from extremely rare regimens. For every remaining treatment hyperedge, we estimate its average probability of HFpEF hospitalization among snapshots in that age group using the same doubly robust procedure as in Section 3, and rank all regimens in ascending order of estimated risk.

Table 2 provides the mapping from numeric treatment IDs to drug-class combinations. Figure 3 visualizes the age-stratified rankings: the left panel shows the ordering for patients older than 65 years, and the right panel shows the ordering for patients 65 years or younger, with bar lengths representing the estimated average hospitalization risk for each regimen.

**Figure 3:**
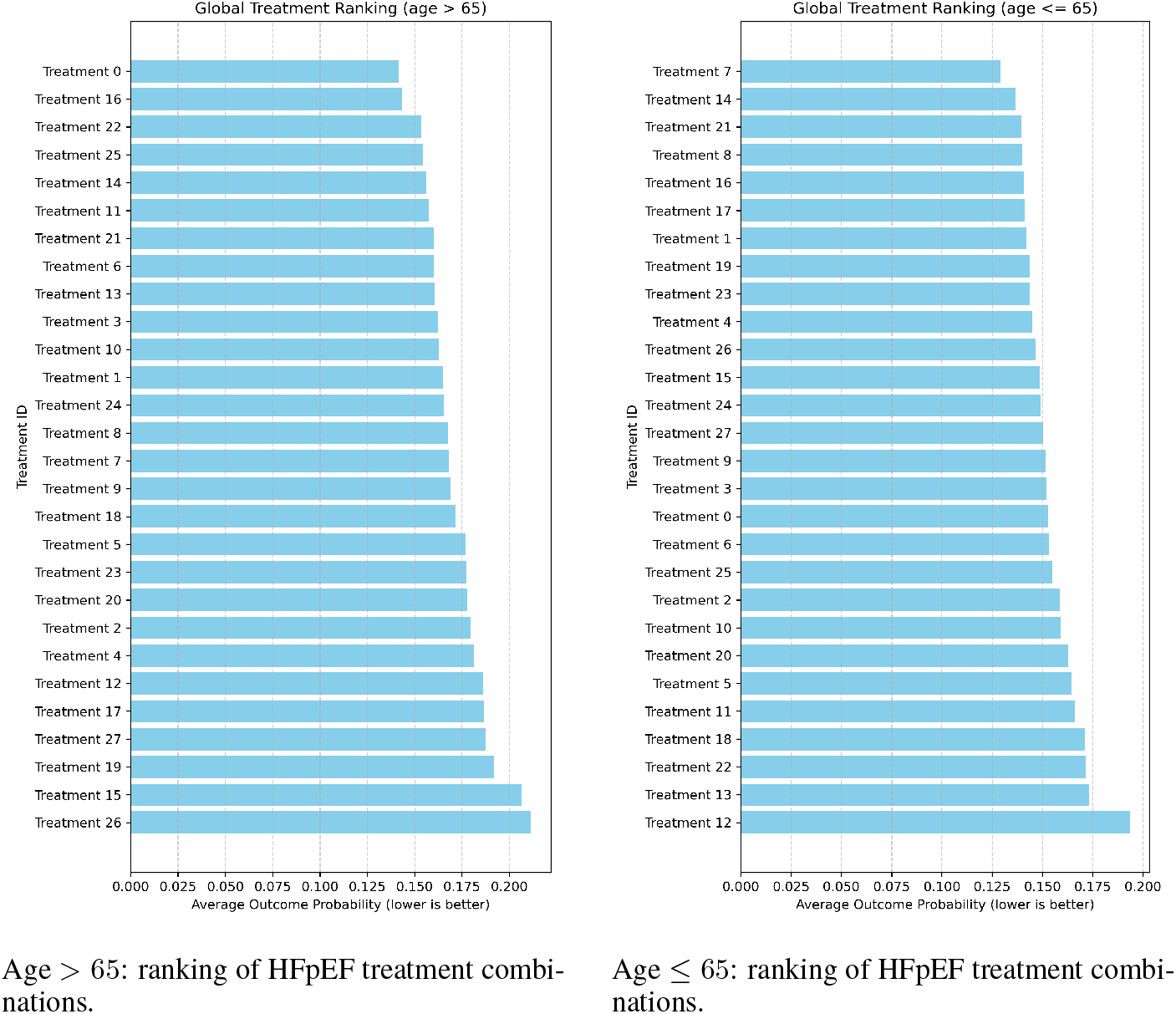
Age-stratified ranking of HFpEF treatment combinations for patients older than 65 years (left) and 65 years or younger (right). Lower bars correspond to lower estimated HFpEF hospitalization risk.

Overall, the ordering of regimens is broadly similar across age groups, with combination therapies that include RAAS inhibition and diuretics generally achieving lower estimated risk than simple monotherapies. However, several treatment IDs shift noticeably between the two rankings (e.g., RAAS monotherapy and SGLT-containing combinations), suggesting that older and younger patients may benefit differently from specific drug combinations. These age-related differences further support the need for subgroup-aware treatment effect estimation in HFpEF.

## 5 Conclusion and Future Work

In this paper, we formulated comparative effectiveness for multi-drug HFpEF treatment as an observational analogue of a randomized trial, and proposed HyperDR, a hypergraph-based doubly robust framework that represents drug classes and their combinations as hyperedges and jointly trains propensity and outcome models with an augmented inverse-probability–weighted objective. On a large HFpEF EHR cohort, HyperDR improves hospitalization risk prediction over baselines and yields interpretable treatment rankings and subgroup-specific patterns.

Future work will extend HyperDR from static combinations to ordered treatment strategies, explicitly modeling the initiation, escalation, and discontinuation sequence of drug classes as dynamic treatment regimes over a hypergraph action space. Additional directions include cross-site validation on heterogeneous health systems, integrating prior RCT and guideline evidence into the hypergraph, and incorporating uncertainty quantification and sensitivity analysis for unmeasured confounding to further strengthen its use in decision support and trial design.

## Data Availability

The data used in this study are not publicly available due to institutional and legal data protection policies.

## References

Connie W Tsao, Aaron W Aday, Zaid I Almarzooq, Cheryl AM Anderson, Pankaj Arora, Christy L Avery, Carissa M Baker-Smith, Andrea Z Beaton, Amelia K Boehme, Alfred E Buxton, et al. Heart disease and stroke statistics—2023 update: a report from the american heart association. Circulation, 147(8):e93–e621, 2023.

Margaret M Redfield. Heart failure with preserved ejection fraction. New England Journal of Medicine, 375(19):1868–1877, 2016.

Paul A Heidenreich, Biykem Bozkurt, David Aguilar, Larry A Allen, Joni J Byun, Monica M Colvin, Anita Deswal, Mark H Drazner, Shannon M Dunlay, Linda R Evers, et al. 2022 aha/acc/hfsa guideline for the management of heart failure: a report of the american college of cardiology/american heart association joint committee on clinical practice guidelines. Journal of the American College of Cardiology, 79(17):e263–e421, 2022.

John JV McMurray and Milton Packer. How should we sequence the treatments for heart failure and a reduced ejection fraction? a redefinition of evidence-based medicine. Circulation, 143(9): 875–877, 2021.

Scott D Solomon, John JV McMurray, Brian Claggett, Rudolf A de Boer, David DeMets, Adrian F Hernandez, Silvio E Inzucchi, Mikhail N Kosiborod, Carolyn SP Lam, Felipe Martinez, et al. Dapagliflozin in heart failure with mildly reduced or preserved ejection fraction. New England Journal of Medicine, 387(12):1089–1098, 2022.

Christian Lienhardt, Sharlette V Cook, Marcos Burgos, Victoria Yorke-Edwards, Leen Rigouts, Gladys Anyo, Sang-Jae Kim, Amina Jindani, Don A Enarson, Andrew J Nunn, et al. Efficacy and safety of a 4-drug fixed-dose combination regimen compared with separate drugs for treatment of pulmonary tuberculosis: the study c randomized controlled trial. Jama, 305(14):1415–1423, 2011.

Miguel A Hernan and James M Robins. Using big data to emulate a target trial when a randomized trial is not available. American journal of epidemiology, 183(8):758–764, 2016.

Fredrik Johansson, Uri Shalit, and David Sontag. Learning representations for counterfactual inference. In International conference on machine learning, pages 3020–3029. PMLR, 2016.

Claudia Shi, David Blei, and Victor Veitch. Adapting neural networks for the estimation of treatment effects. Advances in neural information processing systems, 32, 2019.

Chinmay Belthangady, Stefanos Giampanis, Ivana Jankovic, Will Stedden, Paula Alves, Stephanie Chong, Charlotte Knott, and Beau Norgeot. Causal deep learning reveals the comparative effectiveness of antihyperglycemic treatments in poorly controlled diabetes. Nature Communications, 13(1):6921, 2022.

Ruoqi Liu, Lang Li, and Ping Zhang. Estimating the treatment effects of multiple drug combinations on multiple outcomes in hypertension. Cell Reports Medicine, 6(2), 2025.

Guido W Imbens and Donald B Rubin. Causal inference in statistics, social, and biomedical sciences. Cambridge university press, 2015.

Uri Shalit, Fredrik D Johansson, and David Sontag. Estimating individual treatment effect: generalization bounds and algorithms. In International conference on machine learning, pages 3076–3085. PMLR, 2017.

YiFan Zhang, Hanlin Zhang, Zachary Chase Lipton, Li Erran Li, and Eric Xing. Exploring transformer backbones for heterogeneous treatment effect estimation. Transactions on Machine Learning Research, 2024.

Sonali Parbhoo, Stefan Bauer, and Patrick Schwab. Ncore: Neural counterfactual representation learning for combinations of treatments. arXiv preprint arXiv:2103.11175, 2021.

